# Cervical atrophy following complete thoracic spinal cord injury: Insights from a multinational cohort

**DOI:** 10.1101/2025.11.24.25340899

**Authors:** Yann Quidé, Negin Hesam-Shariati, Zina Trost, Thiago Folly de Campos, William R. Willoughby, Mark Bolding, Rachel E. Cowan, Pauline Zahara, Sylvia M. Gustin

## Abstract

**Background:** Spinal cord injury (SCI) results in neurodegeneration both at and above the lesion site. While cervical cord atrophy is well characterized in populations of mixed cervical, thoracic and/or lumbar injuries, the remote morphological changes in cervical cord following thoracic SCI remain unclear. The present study aimed to quantify cervical spinal cord morphology at C2-C3 in individuals with thoracic SCI and compare these metrics to matched controls.

**Methods:** Participants were 60 adults with chronic thoracic SCI and 60 neurologically healthy controls matched for age and sex. Extracted cervical metrics included mean cross-sectional area (CSA), antero-posterior (AP) and right-left (RL) diameters, eccentricity, solidity, orientation, and cord length. Group differences were assessed using linear mixed models adjusted for age, sex and scanning sites. Impacts of experiencing SCI-related chronic neuropathic pain on these metrics were explored.

**Results:** Compared to controls, the thoracic SCI group showed significantly reduced cervical CSA, along with smaller AP and RL diameters, consistent with remote atrophy. Eccentricity was increased, indicating a more flattened cord profile. Solidity, orientation, and cord length showed no group differences, supporting metric reliability and the absence of segmentation artefacts. Compared to controls, increased eccentricity was driven by people with SCI who experience chronic neuropathic pain. The presence of chronic neuropathic pain did not significantly influence other morphometric outcomes.

**Conclusions:** Thoracic SCI is associated with significant remote cervical cord degeneration, even in the absence of direct cervical injury. Results highlight that neurodegenerative processes propagate along ascending and descending spinal pathways. Cervical morphometry metrics, particularly CSA and eccentricity, may represent biomarkers of distal neurodegeneration following thoracic SCI, and inform future therapeutic strategies.

## Introduction

The latest global burden data highlights a concerning trend: the prevalence of spinal cord injury (SCI) has increased by over 80% between 1990 and 2019, currently affecting around 20.6 million people worldwide (Collaborators 2023). While no precise global data exist, regional studies suggest that approximately 45% of people with spinal cord injury experience paraplegia (Welfare 2024, Center 2025). Thoracic-level injuries are associated with severe long-term impairments in gait, trunk control, autonomic regulation, sexual, bowel and bladder functions, all of which substantially affect quality of life (Lude, Kennedy et al. 2014). Chronic pain affects 50–80% of individuals with SCI and contributes significantly to overall disability (Strom, Manum et al. 2022).

Although the primary injury is distal to the cervical enlargement, Wallerian degeneration, the reduction in the axon segment away from the cell body (distal), and retrograde axonal degeneration, the reduction in the axon segment toward the cell body (proximal), can lead to the atrophy of ascending sensory tracts and descending motor pathways, producing measurable structural changes in cervical segments (Azzarito, Seif et al. 2020). Cervical cord magnetic resonance imaging can identify remote neurodegeneration in individuals with thoracic SCI, without the confounds of local trauma or metal hardware at the lesion site. Rostral cervical morphological changes may provide biomarkers of injury severity and offer potential prognostic value or a target for monitoring neuroprotective interventions.

Imaging-based morphometric studies in SCI have consistently reported cervical spinal cord atrophy (Trolle, Goldberg et al. 2023). However, most studies have used small and mixed cohorts of patients with cervical, thoracic, and/or lumbar SCIs, limiting the ability to disentangle effects specific to lesion level (e.g., Jutzeler, Huber et al. 2016, Wyss, Huber et al. 2019, Azzarito, Seif et al. 2020, Pfyffer, Wyss et al. 2020, David, Pfyffer et al. 2021). This heterogeneity can confound interpretation, as the inclusion of cervical lesions may cause direct local atrophy as well as rostral changes, whereas thoracic lesions are more likely to induce remote degeneration through tract disruption. In addition, studies that have focused primarily on thoracic SCI were of small sample sizes (e.g., Awai and Curt 2015). A systematic review and meta-analysis investigating cervical spinal cord cross-sectional area (CSA) in SCI highlighted that although significant atrophy is evident across studies, injury-level effects vary and are often not isolated, contributing to between-study heterogeneity (Trolle, Goldberg et al. 2023). It thus remains unclear whether cervical cord atrophy is a universal consequence in SCI, independently of the lesion location, or whether the pattern of degeneration differs for thoracic injuries specifically.

The heterogeneity in methodologies used to measure CSA across studies is another limitation to our understanding of the consequences of SCI. Although using T2-weighted imaging of the spinal cord is the gold-standard approach for spinal cord imaging, many projects routinely acquire whole-brain T1-weighted images that can be used, to some extent, to assess cervical cord morphology. The Enhancing NeuroImaging Genetics through Meta-Analysis (ENIGMA) Consortium (Thompson, Jahanshad et al. 2020) has developed the ENIGMA-Cervical spinal cord protocol, a standardized pipeline to quantify cord structure and shape from whole-brain T1-weighted scans. In addition to CSA, key additional metrics can inform on the integrity of the cervical cord including indices of antero-posterior and right-left (transverse) diameters, eccentricity, and solidity, each capturing distinct aspects of cord structure or deformation.

The present study analysed cervical spinal cord morphometry from whole-brain T1-weighted MRI scans in a cohort of individuals with complete thoracic SCI compared with age- and sex-matched non-SCI controls. Participants with complete thoracic SCI were expected to exhibit smaller cervical CSA (at C2 and C3) relative to controls, reflecting remote Wallerian degeneration. In addition, these individuals were expected to show smaller diameters (Jutzeler, Huber et al. 2016) and increased eccentricity, consistent with directional atrophy or morphological remodelling. Finally, exploratory analyses compared these metrics between subgroups of people with SCI who experience chronic neuropathic pain (SCI-NP) and those who did not (SCI-nonNP). Experiencing chronic neuropathic pain was expected to be associated with larger effects of SCI on cervical cord integrity.

## Methods

### Participants

Participants were 60 volunteers with established complete thoracic SCI, defined as having no sensory and motor function in the lowest sacral segments, and 60 age and sex matched healthy, able-bodied controls (HC) without pain or spinal cord injury. All participants were drawn from different cross-sectional studies that used the same clinical and imaging protocols. Neurological level of injury was determined using the American Spinal Injury Association (ASIA) Impairment Scale (AIS) (Kirshblum and Waring 2014), with all participants classified as AIS grade A. Among the 60 SCI participants, 43 experienced persistent neuropathic pain (SCI-NP) below level of injury for more than six months as defined by the International Association for the Study of Pain SCI pain taxonomy (Bryce, Biering-Sorensen et al. 2012, Rosner, de Andrade et al. 2023) and 17 did not experience neuropathic pain (SCI-nonNP). All participants (SCI-NP, SCI-nonNP, HC) gave informed written consent for all procedures. Each sub-study was approved by institutional Human Research Ethics Committees (University of New South Wales (HC15206; HC210932; iRECS4895), the University of Sydney (HREC06287), Northern Sydney Local Health District (1102-066M), University of Alabama at Birmingham Institutional Review Board (IRB-300001463) and Texas A&M University (IRB ID: MOD00000295)).

### Neuroimaging

Whole-brain 3D T1-weighted Magnetization Prepared Rapid Gradient Echo (MPRAGE) scans were acquired for all participants at each site. Details of the sequences used at each site are provided in Supplementary Material. A radiologist reviewed all scans before releasing them to the study investigators, and an additional visual inspection for gross artefacts and movements (presence of excessive ringing that would not allow identification of two adjacent brain regions) was performed.

The ENIGMA-Cervical spinal cord protocol (v1.0; https://github.com/Harding-Lab/enigma-ataxia/tree/master/SpinalCord), developed by the ENIGMA-Ataxia working group, was used to process the cervical spinal cord section from whole-brain T1-weighted scans. This protocol calls the Spinal Cord Toolbox (SCT, v6.5; https://spinalcordtoolbox.com/) (De Leener, Levy et al. 2017) to segment, quantify and quality assess the cervical spinal cord. All images were inspected to ensure that at least C2 vertebral level was covered. Briefly, cervical spinal cords were segmented using an automated deep-learning algorithm (Gros, De Leener et al. 2019), followed by visual inspection of all segmentations before manual correction where necessary. Individual images were then registered to the PAM50 standard template (De Leener, Fonov et al. 2018), and the C2 and C3 vertebral levels were manually marked at the posterior tip of the vertebral discs (Ullmann, Pelletier Paquette et al. 2014, Dupont, De Leener et al. 2017). In some cases, depending on the images field of view or brain size, only C2 was visible. Finally, quantification of the *mean CSA* (in mm^2^) of the cervical spinal cord was performed. In addition to CSA, six other shape metrics were extracted. The *antero-posterior* (AP, in mm) and *right-left* (RL, in mm) *diameters* represent the length of the major and minor axes of the cord. The *eccentricity* of the cord (between 0 and 1) is the ratio of the focal distance to the major axis length. The *solidity* of the cord (between 0 and 1) is the ratio of the CSA to the CSA convex, which can detect strong compression. Finally, the *orientation* is the angular tilt of the cord’s AP axis relative to the image (in degrees), and the *length* of the segmentation (the superior-inferior extent of the segmented cord region) is computed by summing the slice thickness (corrected for the centreline angle at each slice) across the specified superior-inferior region. These last two metrics are primarily used as quality-control metrics to detect registration or acquisition distortions, ensuring that measured differences in CSA or shape are not artefactual.

### Statistical analyses

All statistical analyses were performed using R (v4.4.0; R Core Team 2024) in RStudio (version 2025.05.0+496; Posit team 2025). A series of linear mixed-effect models, using the *lme4* (v1.1.35.4; Bates, Mächler et al. 2015) and *lmerTest* (v3.1.3; Kuznetsova, Brockhoff et al. 2017) packages, were used to compare the spinal cord metrics among the groups (one model per metric); first between the all SCI and HC groups, and second between SCI-NP, SCI-nonNP and HC groups, to determine if the experience of chronic neuropathic pain influenced the effects of SCI on cord metrics. Each model included group as a fixed effect and scanning site as a random effect, with age and sex entered as covariates. False-Discovery Rate (FDR) correction (statistical significance set at *pFDR* < 0.05) using the Benjamini and Hochberg method (Benjamini and Hochberg 1995), was applied to account for the number of metrics (m = 7), separately for each cross-sectional location (C2 or C3). Indices of effect size (*Cohen’s d*) for group differences were extracted from the linear mixed effect models. Exploratory Spearman’s correlations were performed to determine the relationship between age and cross-sectional cord metrics, separately in HC and SCI groups, as well as with the time since injury (in years) in the SCI group only; FDR corrections were applied to account for the number of metrics separately for each cross-sectional location.

## Results

Demographic details are summarized in Table 1. Participants with SCI did not differ significantly from the HC group for age or sex, but group distributions differed across scanning sites. Of the 60 participants with SCI, 43 experienced chronic neuropathic pain following their SCI, and 17 did not.

**Table 1.**
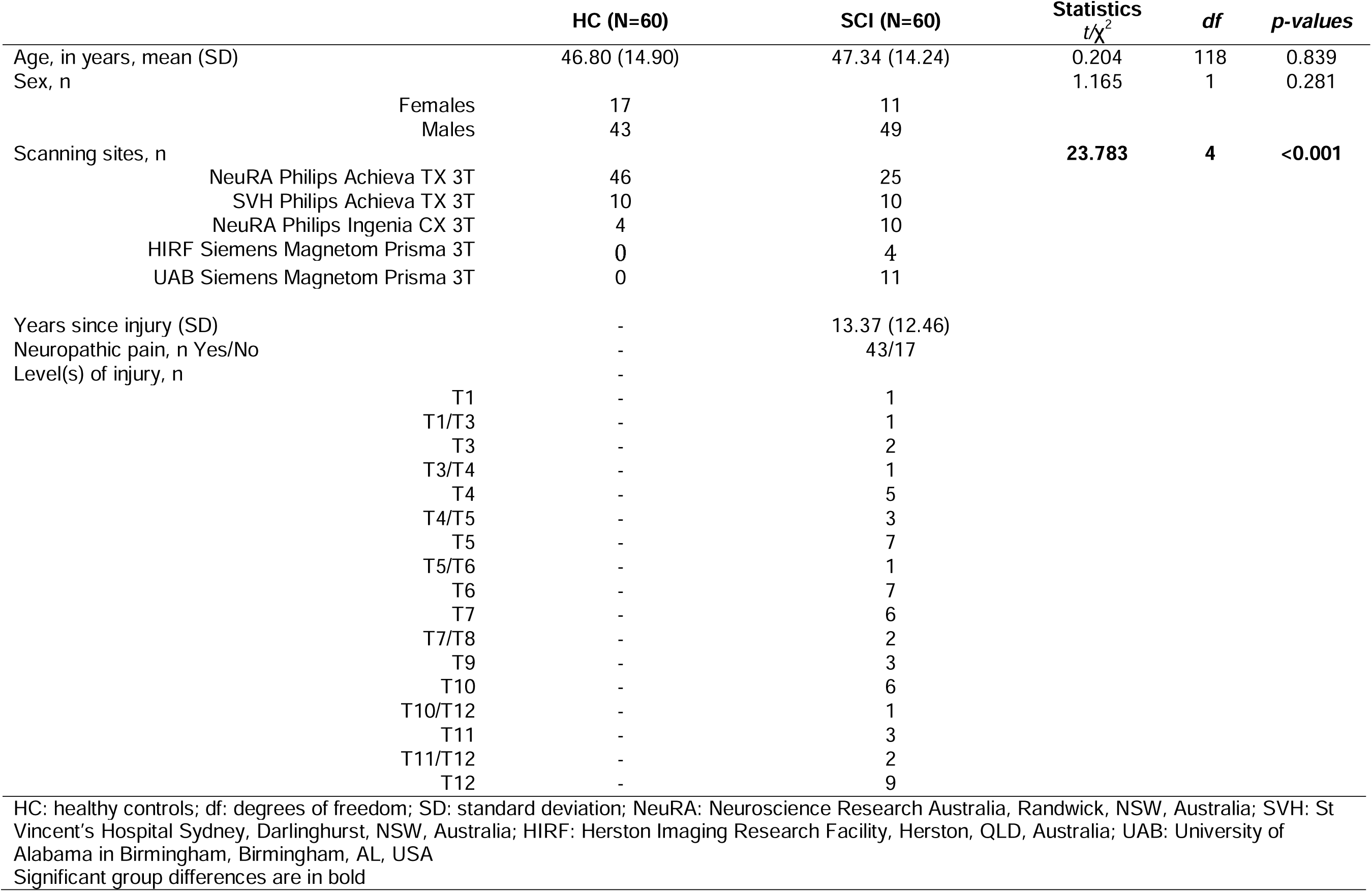
Sociodemographic and clinical characteristics of the studied cohort.

### Cross sectional metrics differences between SCI and HC

Statistical details of the linear mixed effect models are presented Table 2 and illustrated in Figure 1. When comparing all SCI participants to the HC group, having an SCI was significantly (*pFDR* < 0.05) associated with smaller mean CSA (C2: *Cohen’s d =* −1.582; C3: *Cohen’s d =* −1.869), mean AP (C2: *Cohen’s d =* −1.606; C3: *Cohen’s d =* −2.356) and mean RL diameters (C2: *Cohen’s d =* −0.932; C3: *Cohen’s d =* −1.097) as well as larger mean eccentricity (C2: *Cohen’s d =* 0.605; C3: *Cohen’s d =* 0.718). Mean solidity (C2: Cohen’s d = −0.233; C3: Cohen’s d = −0.019), mean orientation (C2: Cohen’s d = 0.120; C3: Cohen’s d = 0.268) and mean length (C2: Cohen’s d = 0.215; C3: Cohen’s d = 0.150) did not significantly differ between the groups (*pFDR* > 0.05).

**Figure 1.**
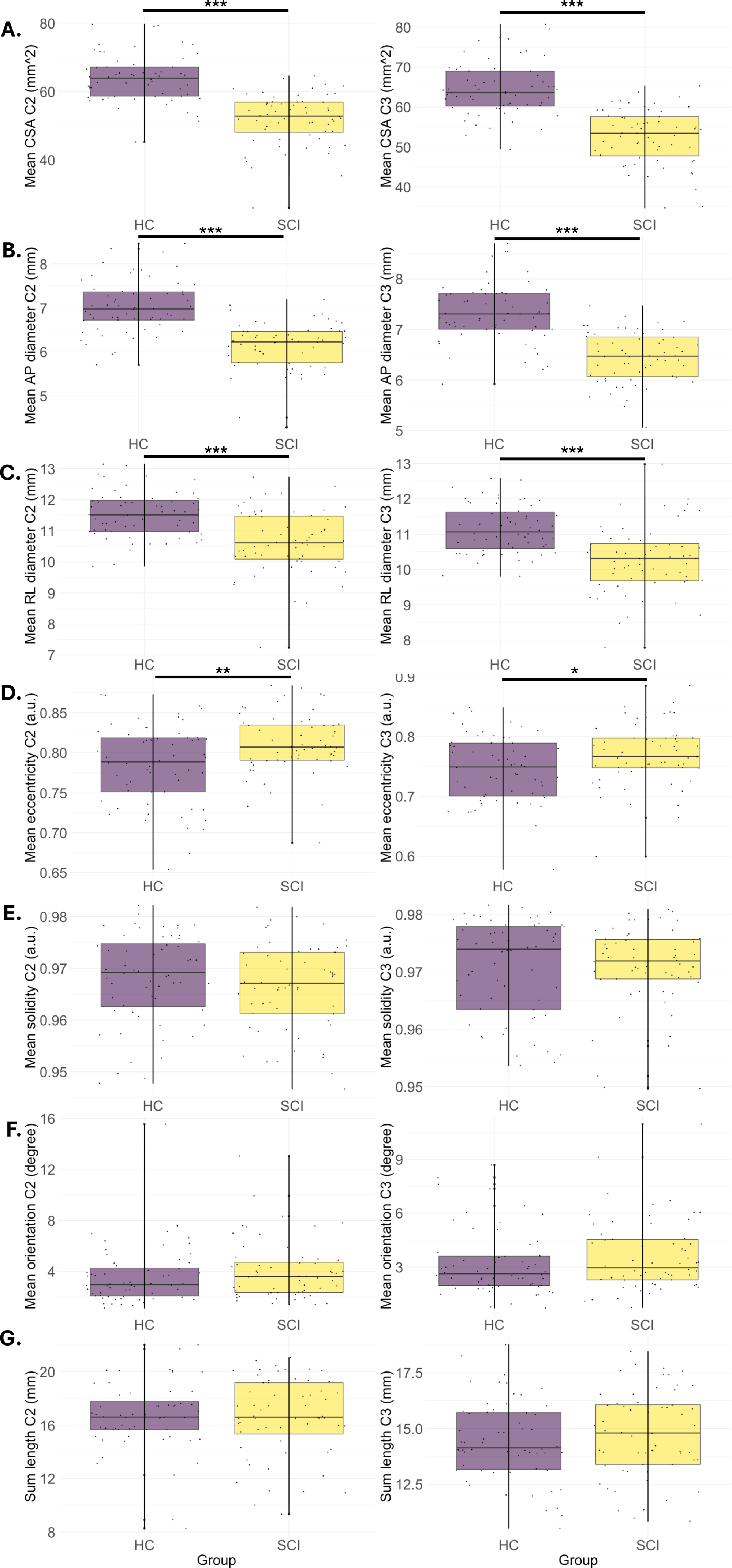
Group differences in cervical cord morphometrics between people with complete thoracic spinal cord injury and controls. Compared to the control group (HC, in purple), the spinal cord injury group (SCI, in yellow) showed significantly smaller (A) cross-sectional area (CSA), (B) antero-posterior (AP) and (C) left-right (RL) diameters, and (D) larger eccentricity of the cervical cord at C2 (left panel) and C3 (right panel). (E) Solidity, (F) orientation and (G) length did not significantly differ between groups. * *p* < 0.05; ** *p* < 0.01; *** *p* < 0.001

**Table 2.** Effects of a thoracic SCI on c-spine morphometrics (C2 and C3 levels)

|  |  |  |  |  |  |  |  |  |
| --- | --- | --- | --- | --- | --- | --- | --- | --- |
| **C2 metric** | **Estimate** | **SE** | **95%LLCI** | **95%ULCI** | **t-value** | **p-value** | **pFDR** | **Cohen's d** |
| Mean CSA | **-11.228** | **1.348** | **-13.870** | **-8.586** | **-8.330** | **2.37E-13** | **1.66E-12** | **-1.582** |
| Mean AP | **-0.845** | **0.108** | **-1.056** | **-0.635** | **-7.858** | **5.73E-12** | **2.01E-11** | **-1.606** |
| Mean RL | **-0.810** | **0.172** | **-1.146** | **-0.473** | **-4.716** | **7.62E-06** | **1.78E-05** | **-0.932** |
| Eccentricity | **0.026** | **0.008** | **0.011** | **0.042** | **3.257** | **0.001** | **0.003** | **0.605** |
| Solidity | -0.002 | 0.002 | -0.005 | 0.001 | -1.248 | 0.214 | 0.290 | -0.233 |
| Orientation | 0.286 | 0.447 | -0.590 | 1.162 | 0.640 | 0.524 | 0.524 | 0.120 |
| Length | 0.575 | 0.496 | -0.397 | 1.547 | 1.159 | 0.249 | 0.290 | 0.215 |
| **C3 metric** | **Estimate** | **SE** | **LLCI** | **ULCI** | **t-value** | **p-value** | **pFDR** | **Cohen's d** |
| Mean CSA | **-12.123** | **1.205** | **-14.484** | **-9.762** | **-10.064** | **1.66E-17** | **1.17E-16** | **-1.869** |
| Mean AP | **-0.870** | **0.100** | **-1.066** | **-0.675** | **-8.721** | **6.08E-12** | **2.13E-11** | **-2.356** |
| Mean RL | **-0.884** | **0.150** | **-1.177** | **-0.591** | **-5.906** | **3.57E-08** | **8.33E-08** | **-1.097** |
| Eccentricity | **0.024** | **0.010** | **0.005** | **0.043** | **2.419** | **0.020** | **0.034** | **0.718** |
| Solidity | -1.49E-04 | 0.001 | -0.003 | 0.003 | -0.104 | 0.917 | 0.917 | -0.019 |
| Orientation | 0.502 | 0.348 | -0.179 | 1.183 | 1.445 | 0.151 | 0.212 | 0.268 |
| Length | 0.232 | 0.329 | -0.412 | 0.876 | 0.705 | 0.483 | 0.563 | 0.150 |

SCI: spinal cord injury; SE: standard error; LLCI/ULCI: lower-level and upper-level 95% confidence interval; FDR: false discovery rate correction; CSA: cross-sectional area; AP: anteroposterior diameter; RL: right-left diameter Group differences surviving FDR correction are in bold

### Cross sectional metrics differences between SCI-NP, SCI-nonNP and HC

Statistical details of the linear mixed effect models comparing the three groups are presented Table 3 and illustrated in Figure 2. Compared to the control group, both SCI-NP and SCI-nonNP showed significantly smaller mean CSA (SCI-NP: C2: *Cohen’s d =* −1.334, C3: *Cohen’s d =* −1.650; SCI-nonNP: C2: *Cohen’s d =* −1.245, C3: *Cohen’s d =* −1.369), mean AP (SCI-NP: C2: *Cohen’s d =* −1.740, C3: *Cohen’s d =* −2.999; SCI-nonNP: C2: *Cohen’s d =* −1.072; C3: *Cohen’s d =* −1.118) and mean RL diameters (SCI-NP: C2: *Cohen’s d =* −0.793, C3: *Cohen’s d =* −0.909; SCI-nonNP: C2: *Cohen’s d =* −0.727, C3: *Cohen’s d =* −0.925). Mean eccentricity was larger in the SCI-NP group (C2: *Cohen’s d =* 0.570, C3: *Cohen’s d =* 0.476), but not in the SCI-nonNP group (C2: *Cohen’s d =* 0.373, C3: *Cohen’s d =* 0.202). Mean solidity (SCI-NP: C2: Cohen’s d = −0.238, C3: Cohen’s d = −0.010; SCI-nonNP: C2: Cohen’s d = −0.143, C3: Cohen’s d = −0.027), orientation (SCI-NP: C2: Cohen’s d = 0.169, C3: Cohen’s d = 0.252; SCI-nonNP: C2: Cohen’s d = 0.029, C3: Cohen’s d = 0.166), and mean length (SCI-NP: C2: Cohen’s d = 0.215, C3: Cohen’s d = 0.259; SCI-nonNP: C2: Cohen’s d = 0.124, C3: Cohen’s d = 0.018) did not significantly differ between the groups (*pFDR* > 0.05). The two SCI groups did not significantly differ from each other on any of the cervical cord metrics studied (see details Table 3).

**Figure 2.**
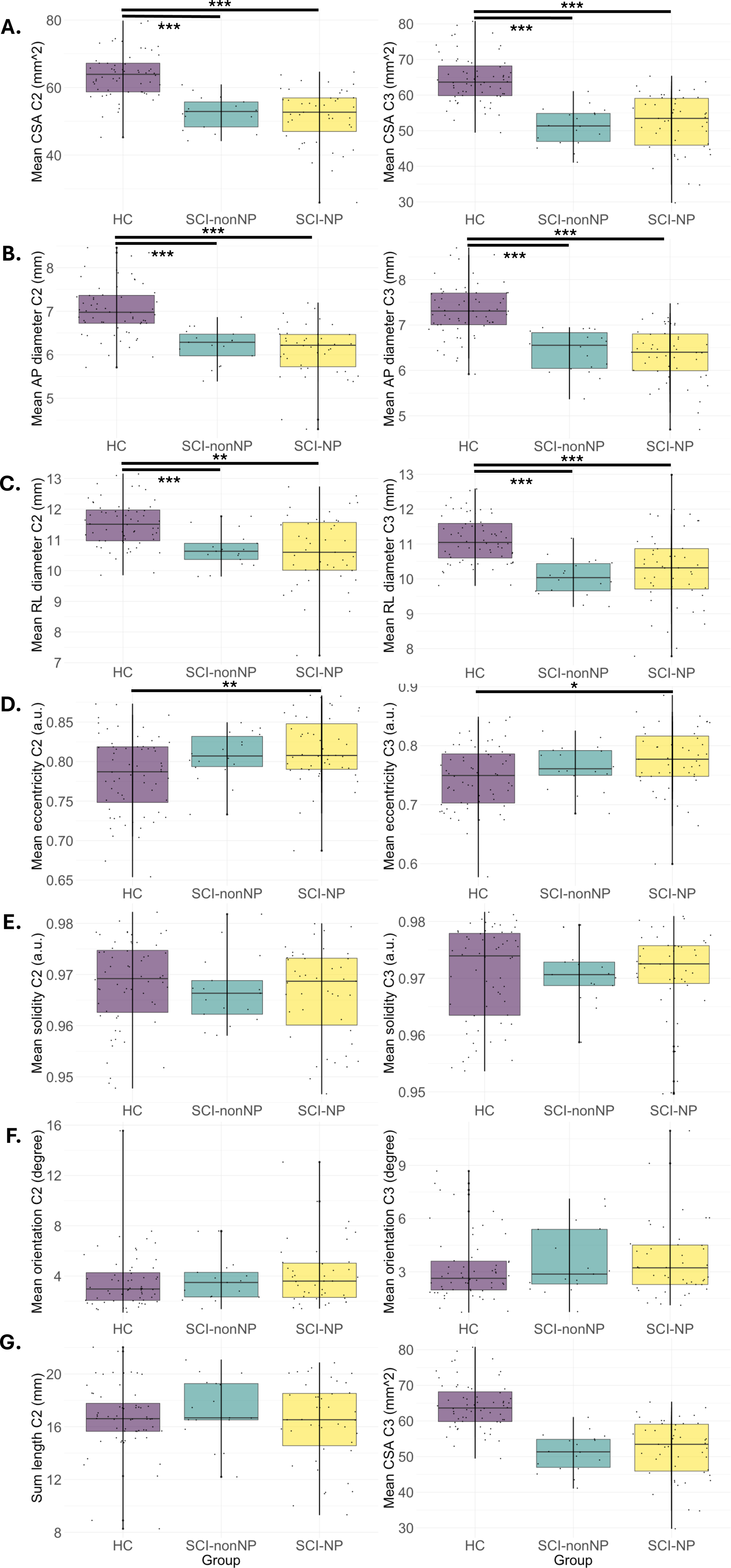
Group differences in cervical cord morphometrics between people with complete thoracic spinal cord injury with or without chronic neuropathic pain, and controls. Compared to the control group (HC, in purple), the spinal cord injury group without neuropathic pain (SCI-nonNP, in green), and the SCI group with neuropathic pain (SCI-NP, in yellow) showed significantly smaller (A) cross-sectional area (CSA), (B) antero-posterior (AP) and (C) left-right (RL) diameters of the cervical cord at C2 (left panel) and C3 (right panel). (D) Significant larger eccentricity was only evident in the SCI-NP group relative to control. (E) Solidity, (F) orientation and (G) length did not significantly differ between groups. * *p* < 0.05; ** *p* < 0.01; *** *p* < 0.001

**Table 3.** Effects of a thoracic SCI on c-spine morphometrics (C2 and C3 levels), among people who experienced or not chronic neuropathic pain.

| **C2 metric** | **Estimate** | **SE** | **95%LLCI** | **95%ULCI** | **t-value** | **p-value** | **pFDR** | **Cohen's d** |
| --- | --- | --- | --- | --- | --- | --- | --- | --- |
| ***SCI-NP vs HC*** |  |  |  |  |  |  |  |  |
| Mean CSA | **-10.297** | **1.593** | **-13.420** | **-7.174** | **-6.462** | **4.57E-09** | **3.199E-08** | **-1.334** |
| Mean AP | **-0.820** | **0.125** | **-1.065** | **-0.575** | **-6.557** | **1.75E-08** | **6.125E-08** | **-1.740** |
| Mean RL | **-0.708** | **0.202** | **-1.103** | **-0.312** | **-3.504** | **7.628E-04** | **0.002** | **-0.793** |
| Eccentricity | **0.027** | **0.009** | **0.010** | **0.045** | **3.054** | **0.003** | **0.005** | **0.570** |
| Solidity | -0.002 | 0.002 | -0.006 | 0.002 | -1.168 | 0.246 | 0.299 | -0.238 |
| Orientation | 0.408 | 0.525 | -0.622 | 1.438 | 0.777 | 0.439 | 0.439 | 0.169 |
| Length | 0.680 | 0.596 | -0.487 | 1.847 | 1.142 | 0.256 | 0.299 | 0.215 |
| ***SCI-nonNP vs HC*** | **Estimate** | **SE** | **95%LLCI** | **95%ULCI** | **t-value** | **p-value** | **pFDR** | **Cohen's d** |
| Mean CSA | **-12.352** | **1.874** | **-16.024** | **-8.679** | **-6.592** | **1.47E-09** | **1.029E-08** | **-1.245** |
| Mean AP | **-0.870** | **0.153** | **-1.169** | **-0.570** | **-5.694** | **9.96E-08** | **3.486E-07** | **-1.072** |
| Mean RL | **-0.929** | **0.241** | **-1.402** | **-0.456** | **-3.851** | **1.962E-04** | **4.578E-04** | **-0.727** |
| Eccentricity | 0.024 | 0.012 | 0.000 | 0.048 | 1.998 | 0.048 | 0.084 | 0.373 |
| Solidity | -0.002 | 0.002 | -0.006 | 0.003 | -0.754 | 0.453 | 0.601 | -0.143 |
| Orientation | 0.096 | 0.623 | -1.124 | 1.316 | 0.154 | 0.878 | 0.878 | 0.029 |
| Length | 0.445 | 0.681 | -0.889 | 1.779 | 0.654 | 0.515 | 0.601 | 0.124 |
| ***SCI-NP vs SCI-nonNP*** | **Estimate** | **SE** | **95%LLCI** | **95%ULCI** | **t-value** | **p-value** | **pFDR** | **Cohen's d** |
| Mean CSA | 2.055 | 2.134 | -2.128 | 6.237 | 0.963 | 0.338 | 0.830 | 0.187 |
| Mean AP | 0.050 | 0.169 | -0.282 | 0.382 | 0.295 | 0.769 | 0.830 | 0.065 |
| Mean RL | 0.221 | 0.272 | -0.311 | 0.754 | 0.814 | 0.418 | 0.830 | 0.165 |
| Eccentricity | 0.003 | 0.013 | -0.022 | 0.028 | 0.217 | 0.828 | 0.830 | 0.041 |
| Solidity | -0.001 | 0.003 | -0.006 | 0.005 | -0.215 | 0.830 | 0.830 | -0.041 |
| Orientation | 0.312 | 0.705 | -1.070 | 1.695 | 0.443 | 0.659 | 0.830 | 0.088 |
| Length | 0.235 | 0.791 | -1.315 | 1.785 | 0.297 | 0.767 | 0.830 | 0.055 |
| **C3 metric** | **Estimate** | **SE** | **95%LLCI** | **95%ULCI** | **t-value** | **p-value** | **pFDR** | **Cohen's d** |
| ***SCI-NP vs HC*** |  |  |  |  |  |  |  |  |
| Mean CSA | **-11.661** | **1.318** | **-14.245** | **-9.077** | **-8.845** | **1.27E-14** | **8.89E-14** | **-1.650** |
| Mean AP | **-0.857** | **0.113** | **-1.078** | **-0.635** | **-7.587** | **5.20E-08** | **1.82E-07** | **-2.999** |
| Mean RL | **-0.794** | **0.163** | **-1.114** | **-0.475** | **-4.873** | **3.55E-06** | **8.28E-06** | **-0.909** |
| Eccentricity | **0.027** | **0.011** | **0.006** | **0.049** | **2.554** | **0.012** | **0.021** | **0.476** |
| Solidity | -8.57E-05 | 0.002 | -0.003 | 0.003 | -0.055 | 0.957 | 0.957 | -0.010 |
| Orientation | 0.516 | 0.382 | -0.232 | 1.264 | 1.351 | 0.179 | 0.251 | 0.252 |
| Length | 0.338 | 0.377 | -0.402 | 1.078 | 0.896 | 0.375 | 0.438 | 0.259 |
| **SCI-nonNP vs HC** | **Estimate** | **SE** | **95%LLCI** | **95%ULCI** | **t-value** | **p-value** | **pFDR** | **Cohen's d** |
| Mean CSA | **-13.294** | **1.811** | **-16.842** | **-9.745** | **-7.342** | **3.24E-11** | **2.27E-10** | **-1.369** |
| Mean AP | **-0.880** | **0.147** | **-1.167** | **-0.592** | **-5.988** | **2.49E-08** | **8.72E-08** | **-1.118** |
| Mean RL | **-1.111** | **0.224** | **-1.550** | **-0.672** | **-4.962** | **2.44E-06** | **5.69E-06** | **-0.925** |
| Eccentricity | 0.016 | 0.015 | -0.013 | 0.045 | 1.082 | 0.281 | 0.492 | 0.202 |
| Solidity | -3.09E-04 | 0.002 | -0.005 | 0.004 | -0.143 | 0.887 | 0.925 | -0.027 |
| Orientation | 0.468 | 0.524 | -0.560 | 1.495 | 0.892 | 0.374 | 0.524 | 0.166 |
| Length | 0.044 | 0.468 | -0.873 | 0.962 | 0.095 | 0.925 | 0.925 | 0.018 |
| **SCI-NP vs SCI-nonNP** | **Estimate** | **SE** | **95%LLCI** | **95%ULCI** | **t-value** | **p-value** | **pFDR** | **Cohen's d** |
| Mean CSA | 1.633 | 1.883 | -2.059 | 5.324 | 0.867 | 0.388 | 0.930 | 0.162 |
| Mean AP | 0.023 | 0.157 | -0.285 | 0.330 | 0.145 | 0.885 | 0.930 | 0.038 |
| Mean RL | 0.317 | 0.233 | -0.140 | 0.773 | 1.359 | 0.177 | 0.930 | 0.253 |
| Eccentricity | 0.011 | 0.015 | -0.019 | 0.042 | 0.747 | 0.456 | 0.930 | 0.139 |
| Solidity | 2.23E-04 | 0.002 | -0.004 | 0.005 | 0.099 | 0.921 | 0.930 | 0.019 |
| Orientation | 0.048 | 0.545 | -1.021 | 1.117 | 0.088 | 0.930 | 0.930 | 0.016 |
| Length | 0.294 | 0.514 | -0.714 | 1.302 | 0.571 | 0.570 | 0.930 | 0.131 |

SCI: spinal cord injury; SCI-NP: people with SCI and chronic neuropathic pain; SCI-nonNP: people with SCI and without chronic neuropathic pain; SE: standard error; LLCI/ULCI: lower-level and upper-level 95% confidence interval; FDR: false discovery rate correction; CSA: cross-sectional area; AP: anteroposterior diameter; RL: right-left diameter
Group differences surviving FDR correction are in bold

### Exploratory correlation analyses

Statistical details of the correlation analyses are presented Supplementary Tables S2-S3. Increasing age was significantly associated with smaller C2 (*r* = −0.371, *pFDR* = 0.024) and C3 (*r* = −0.354, *pFDR* = 0.039) mean CSA, as well as with smaller AP diameter (*r* = −0.334, *pFDR* = 0.032) in controls only (see Table S2). No other correlation with age for any other metric or group survived FDR correction. In addition, none of the correlations between C2 or C3 metrics and time since injury survived FDR correction (see Table S3).

## Discussion

This study identified the distal impacts of a complete thoracic SCI on the integrity of the cervical cord. Results indicate that thoracic SCI is associated with significant reductions in cross-sectional area (CSA), antero-posterior and right-left diameters, as well as increased cord eccentricity at both C2 and C3 levels. In contrast, solidity, orientation, or cord length were not different between groups. Compared to able-bodied controls, SCI-NP people had larger cervical cord eccentricity, that was not evident in those with SCI-nonNP.

Consistent with previous studies, smaller cervical CSA was evident in people with thoracic SCI (Trolle, Goldberg et al. 2023). Reduced CSA in the cervical cord likely reflects ascending and descending tract degeneration, including Wallerian degeneration of corticospinal and ascending sensory pathways (Grabher, Callaghan et al. 2015, David, Pfyffer et al. 2021, Fischer, Stern et al. 2021, Schading, David et al. 2023). These results extend those of previous studies (Trolle, Goldberg et al. 2023) and indicate that these degenerative effects are also detectable in individuals with exclusively thoracic SCI, in absence of cervical or lumbar lesions, reinforcing the sensitivity of cervical morphometry to spinal cord pathology even when the lesion is distal.

Both antero-posterior and right-left diameters were significantly reduced in participants with thoracic SCI. Reduced antero-posterior diameter may reflect loss or remodelling of tracts oriented in the antero-posterior plane (e.g., corticospinal or spinothalamic tracts), consistent with a previous study (Jutzeler, Huber et al. 2016). On the other hand, unlike results from that same study (Jutzeler, Huber et al. 2016), reduced right-left diameter was evident in thoracic SCI. Smaller right-left diameter may reflect degeneration of dorsal columns or lateral corticospinal tracts, pointing to asymmetrical degeneration or atrophy. These changes in both antero-posterior and right-left directions suggest generalized cord shrinkage rather than preferential atrophy in one axis, as noted in some studies where injury-level heterogeneity may have introduced variability (Grabher, Callaghan et al. 2015).

People with complete thoracic SCI also showed increased cervical cord eccentricity (more elongated ellipse), indicating a potential subtle shift toward a more flattened cord cross-section; this effect was driven by the experience of chronic neuropathic pain. Increased eccentricity of the cervical cord may indicate shape deformation, potentially reflecting white or grey matter shrinkage in thoracic SCI. Increased eccentricity may reflect mechanical compression or morphological remodelling (Whyte, Melnyk et al. 2020), that can arise from differential degeneration of dorsal columns relative to lateral and ventral tracts. In this context, chronic neuropathic pain may exacerbate cervical cord degeneration, potentially reflecting pain-related stress on the cord. Future studies with larger samples of individuals with SCI who do not experience chronic neuropathic pain are warranted to clarify this differential increase in cervical cord eccentricity.

Solidity of the cervical cord did not differ between groups, suggesting that the internal contour complexity of the cord cross-section remains relatively stable after injury. The absence of group differences in orientation and length metrics supports the reliability of our morphometric measures. Stability of these metrics also indicate that the observed group differences in CSA, diameters and eccentricity reflect genuine neuroanatomical changes rather than registration or segmentation biases.

The presence of cervical morphometric abnormalities in individuals with complete thoracic SCI indicate that neurodegeneration can arise in distal regions of the cord, beyond the level of injury. Remote cervical atrophy has been consistently associated with disruptions in both ascending sensory and descending motor pathways (Freund, Weiskopf et al. 2013, Grabher, Callaghan et al. 2015). Smaller cervical CSA has been associated with impairments in motor recovery, hand function, and sensory deficits, suggesting that morphological metrics in the cervical cord could be used as biomarkers of disease severity and functional prognosis (Lundell, Barthelemy et al. 2011, Huber, David et al. 2018). Distal morphological changes can arise from processes that drive progressive tissue loss, including Wallerian degeneration of axons, axonal disconnection and subsequent demyelination, and persistent inflammation (Freund, Weiskopf et al. 2013, Ahuja, Wilson et al. 2017, David, Mohammadi et al. 2019). The present results highlight the important role of cervical morphometry to reflect remote neurodegenerative changes.

Unlike a previous smaller study (Jutzeler, Huber et al. 2016), the experience of chronic neuropathic pain did not further impact the cervical cord CSA but drove the increase in cord eccentricity in our thoracic SCI group. While pain-related changes have been reported within the thalamocortical systems (Gustin, Wrigley et al. 2014), our findings suggest that structural changes in the cervical cord may progress differently with the experience of neuropathic pain. It is possible that the macrostructural metrics used here, such as CSA as a proxy for quantification of cord tissue remaining, and eccentricity as a proxy for distribution/deformation of the cord, are differentially sensitive to the central sensitization processes implicated in chronic neuropathic pain. Studies including more people with SCI who do not experience chronic neuropathic pain are necessary to confirm this speculative interpretation.

A key strength of this work lies in the inclusion of the largest complete thoracic SCI cohort to date (n = 60), matched to non-SCI controls. Prior studies have often pooled participants across lesion levels (e.g., cervical, thoracic, lumbar), confounding the interpretation of remote atrophy (Trolle, Goldberg et al. 2023). The use of standardized metrics ensures reproducibility and facilitates integration into larger consortia-based efforts. However, this study has several limitations. First, the cross-sectional nature of our data prevents conclusions about the temporal dynamics of atrophy, and the absence of direct functional correlations. We also note that, although not associated with variations in cervical cord metrics (see Supplementary Table S3), time since injury varied from within a year to over 57 years. Future longitudinal studies combining cervical morphometry, diffusion imaging, and clinical/functional outcomes will be critical to delineate the trajectory and clinical significance of remote cord degeneration after thoracic injury. Second, despite the strict selection of people with complete thoracic SCIs, the heterogeneity in the exact lesion locations (T1 to T12) may still confound the results. When possible, future studies of specific levels of injury (e.g., for instance uniquely located at T5) will inform on the extent to which the distance from injury impacts the neurodegenerative effects of thoracic SCI on cervical cord integrity. Third, medication usage was not included in the analysis, as the able-bodied controls were not taking these medications, preventing a direct comparison. Some pharmacological agents commonly used following SCI, such as anti-spasticity drugs or neuropathic pain medications, may influence neural tissue integrity or neuroinflammatory processes, potentially confounding morphometric measures. Although their impact on cervical cord morphology remains poorly understood, future studies should account for medication profiles to better isolate injury-related neurodegeneration from treatment effects.

In conclusion, this study provides robust evidence for distal cervical cord atrophy and altered shape in individuals with complete thoracic SCI. These findings highlight the sensitivity of cervical morphometric measures, particularly CSA, antero-posterior/right-left diameters, and eccentricity, as potential biomarkers of remote spinal degeneration following thoracic SCI. Our results reinforce the importance of incorporating cervical morphometry in SCI research and clinical trials, even when the lesion is anatomically distant from the cervical cord. These metrics have the potential for use in clinical settings for stratifying patients, monitoring progression, and informing future interventions that can preserve or restore spinal cord integrity.

## Data Availability

All data produced in the present study are available upon reasonable request to the authors

## Acknowledgements

This work was supported by a grant from the US Department of Defence (DoD SC190117/Award number: W81XWH-20-1-0775), a project grant from the National Health and Medical Research Council of Australia (ID1084240), a grant from the New South Wales (NSW) Ministry of Health via the Spinal Cord Injury Research Grants Program 2019/20, and a grant from the Wings for Life Spinal Cord Research Foundation. S.M.G. was funded by a Rebecca Cooper Fellowship from the Rebecca L. Cooper Medical Research Foundation. The funding bodies are not involved in the analyses, interpretations or the writing of the findings for publication. We thank all participants for their time and commitment at all stages of the different sub-studies.

