## Supplementary Material for "Cervical atrophy following complete thoracic spinal cord injury: Insights from a multinational cohort"

Yann Quidé^1,2,*^, Negin Hesam-Shariati^1,2^, Zina Trost^3^, Thiago Folly de Campos^1,2^, William R. Willoughby^4^, Mark Bolding^4^, Rachel E. Cowan^5^, Pauline Zahara^1,2^, Sylvia M. Gustin ^1,2^

^1^ NeuroRecovery Research Hub, School of Psychology, The University of New South Wales (UNSW) Sydney, Sydney, NSW, Australia

^2^ Centre for Pain IMPACT, Neuroscience Research Australia, Randwick, NSW, Australia

^3^ Department of Psychological and Brain Sciences, Texas A&M University, College Station, TX, USA

^4^ Department of Radiology, University of Alabama at Birmingham, Birmingham, AL, USA

^5^ Department of Physical Medicine & Rehabilitation, University of Alabama at Birmingham, Birmingham, AL, USA

*** Corresponding author**

Dr Yann Quidé, NeuroRecovery Research Hub, School of Psychology, Biological Sciences (Biolink) building, Level 1, UNSW Sydney, NSW, 2052, Australia.

| **Supplementary Table 1. Scanning parameters at all scanning sites** | | | |
| --- | --- | --- | --- |
| Scanning site | Scanner type | Sequence | Slice orientation |
| NeuRA1 | Philips Achieva TX 3T | 3D T1-weighted Magnetization Prepared Rapid Gradient Echo (MPRAGE) scans; TR/TE = 5.6 ms/2.5 ms, field of view = 250 × 250 × 174 mm, matrix 288 × 288, 200 slices, flip angle = 8°, voxel size 0.9 × 0.9 × 0.9 mm. 16 channels head coil | Sagittal |
| SVH | Philips Achieva TX 3T | 3D T1-weighted Magnetization Prepared Rapid Gradient Echo (MPRAGE) scans; TR/TE = 5.6 ms/2.5 ms, field of view = 250 × 250 × 174 mm, matrix 288 × 288, 200 slices, flip angle = 8°, voxel size 0.9 × 0.9 × 0.9 mm. 16 channels head coil | Sagittal |
| NeuRA2 | Philips Ingenia CX 3T | 3D T1-weighted Magnetization Prepared Rapid Gradient Echo (MPRAGE) scans; TR/TE = 8.0 ms/3.8 ms, field of view = 240 × 240 × 190 mm, matrix 268 × 268, 211 slices, flip angle = 8°, voxel size 0.9 × 0.9 × 0.9 mm. 32 channels head coil | Sagittal |
| HIRF | Siemens MAGNETOM Prisma 3T | 3D T1-weighted Magnetization Prepared Rapid Gradient Echo (MPRAGE) scans; TR/TE = 2400.0 ms/2.22 ms, field of view = 240 × 240 × 188 mm, matrix 288 × 288, 208 slices, flip angle = 8°, voxel size 0.8 × 0.8 × 0.9 mm. 64 channels head-neck coil | Sagittal |
| UAB | Siemens MAGNETOM Prisma 3T | 3D T1-weighted Magnetization Prepared Rapid Gradient Echo (MPRAGE) scans; TR/TE = 2400.0 ms/2.22 ms, field of view = 240 × 240 × 188 mm, matrix 288 × 288, 208 slices, flip angle = 8°, voxel size 0.8 × 0.8 × 0.9 mm. 64 channels head-neck coil | Sagittal |
| NeuRA: Neuroscience Research Australia, Randwick, NSW, Australia; SVH: St Vincent’s Hospital, Sydney, NSW, Australia; HIRF: Herston Imaging Research Facility, Herston, QLD, Australia; UAB: University of Alabama in Birmingham, Birmingham, AL, USA; TR: repetition time; TE: echo time | | | |

| **Table S2. Correlations between age and cord metrics in the healthy able-bodied control and spinal cord injury groups separately** | | | | |
| --- | --- | --- | --- | --- |
| **Group** | **Metric** | **r** | **p-value** | **pFDR** |
| HC | **C2 mean CSA** | **-0.371** | **0.003** | **0.024** |
|  | **C2 mean AP** | **-0.334** | **0.009** | **0.032** |
|  | C2 mean RL | -0.193 | 0.139 | 0.324 |
|  | C2 eccentricity | 0.154 | 0.239 | 0.418 |
|  | C2 solidity | -0.075 | 0.571 | 0.758 |
|  | C2 orientation | -0.060 | 0.649 | 0.758 |
|  | C2 length | 0.001 | 0.996 | 0.996 |
|  | **C3 mean CSA** | **-0.354** | **0.006** | **0.039** |
|  | C3 mean AP | -0.308 | 0.016 | 0.058 |
|  | C3 mean RL | -0.233 | 0.074 | 0.172 |
|  | C3 eccentricity | 0.121 | 0.358 | 0.408 |
|  | C3 solidity | 0.128 | 0.329 | 0.408 |
|  | C3 orientation | -0.173 | 0.185 | 0.324 |
|  | C3 length | -0.109 | 0.408 | 0.408 |
| SCI | C2 mean CSA | -0.174 | 0.183 | 0.598 |
|  | C2 mean AP | -0.145 | 0.268 | 0.598 |
|  | C2 mean RL | -0.125 | 0.342 | 0.598 |
|  | C2 eccentricity | 0.033 | 0.803 | 0.933 |
|  | C2 solidity | -0.131 | 0.320 | 0.933 |
|  | C2 orientation | 0.011 | 0.933 | 0.598 |
|  | C2 length | 0.088 | 0.505 | 0.707 |
|  | C3 mean CSA | -0.143 | 0.275 | 0.445 |
|  | C3 mean AP | -0.084 | 0.524 | 0.611 |
|  | C3 mean RL | -0.131 | 0.318 | 0.445 |
|  | C3 eccentricity | -0.009 | 0.944 | 0.944 |
|  | C3 solidity | -0.269 | 0.037 | 0.262 |
|  | C3 orientation | 0.187 | 0.152 | 0.355 |
|  | C3 length | -0.193 | 0.140 | 0.355 |
| *r*: Pearson correlation coefficient; FDR: false discovery rate correction; HC: healthy able-bodied controls; SCI: spinal cord injury; CSA: cross sectional area; AP: antero-posterior diameter; RL: right-left diameter  Correlations surviving FDR correction are in bold | | | | |

| **Table S3. Correlations between time since injury (in years) and cord metrics in the spinal cord injury group only** | | | |
| --- | --- | --- | --- |
| **Metric** | **r** | **p-value** | **pFDR** |
| C2 mean CSA | -0.101 | 0.442 | 0.516 |
| C2 mean AP | -0.005 | 0.968 | 0.968 |
| C2 mean RL | -0.143 | 0.276 | 0.516 |
| C2 eccentricity | -0.132 | 0.314 | 0.516 |
| C2 solidity | -0.103 | 0.436 | 0.516 |
| C2 orientation | 0.112 | 0.394 | 0.516 |
| C2 length | 0.167 | 0.202 | 0.516 |
| C3 mean CSA | -0.217 | 0.095 | 0.167 |
| C3 mean AP | -0.027 | 0.840 | 0.936 |
| C3 mean RL | -0.302 | 0.019 | 0.134 |
| C3 eccentricity | -0.252 | 0.052 | 0.167 |
| C3 solidity | -0.011 | 0.936 | 0.936 |
| C3 orientation | 0.225 | 0.083 | 0.167 |
| C3 length | 0.083 | 0.530 | 0.742 |
| *r*: Pearson correlation coefficient; FDR: false discovery rate correction; CSA: cross sectional area; AP: antero-posterior diameter; RL: right-left diameter  Correlations surviving FDR correction are in bold | | | |
